# Assessing the impact of non-pharmaceutical interventions (NPI) on the dynamics of COVID-19: A mathematical modelling study in the case of Ethiopia

**DOI:** 10.1101/2020.11.16.20231746

**Authors:** Bedilu Alamirie Ejigu, Manalebish Debalike Asfaw, Lisa Cavalerie, Tilahun Abebaw, Mark Nanyingi, Matthew Baylis

## Abstract

The World Health Organisation (WHO) declared COVID-19 a pandemic on March 11, 2020 and by November 14, 2020 there were 53.3M confirmed cases and 1.3M reported deaths in the world. In the same period, Ethiopia reported 102K cases and 1.5K deaths. Effective public health preparedness and response to COVID-19 requires timely projections of the time and size of the peak of the outbreak. Currently, in the absence of vaccine or effective treatment, the implementation of NPIs (non-pharmaceutical interventions), like hand washing, wearing face coverings or social distancing, is recommended by WHO to bring the pandemic under control. This study proposes a modified Susceptible Exposed Infected and Recovered (SEIR) model to predict the number of COVID-19 cases at different stages of the disease under the implementation of NPIs with different adherence levels in both urban and rural settings of Ethiopia. To estimate the number of cases and their peak time, 30 different scenarios were simulated. The results reveal that the peak time of the pandemic is different in urban and rural populations of Ethiopia. In the urban population, under moderate implementation of three NPIs the pandemic will be expected to reach its peak in December, 2020 with 147,972 cases, of which 18,100 are symptomatic and 957 will require admission to an Intensive Care Unit (ICU). Among the implemented NPIs, increasing the coverage of wearing masks by 10% could reduce the number of new cases on average by one-fifth in urban-populations. Varying the coverage of wearing masks in rural populations minimally reduces the number of cases. In conclusion, the projection result reveals that the projected number of hospital cases is higher than the Ethiopian health system capacity during the peak time. To contain symptomatic and ICU cases within health system capacity, the government should give attention to the strict implementation of the existing NPIs or impose additional public health measures.

## 1 Introduction

The novel Severe Acute Respiratory Syndrome Coronavirus 2 (SARS-CoV-2) was first discovered in Wuhan, China in December 2019 WHO (2020). The associated disease, COVID-19 was declared as a pandemic by the World Health Organisation (WHO) on March 11, 2020. It has led to over 53.5 million confirmed cases and 1.3 million deaths across the globe by November 14, 2020 WHO (2020). In Africa, the burden and impacts of the pandemic are less compared with Europe and the USA. This may be attributed to late arrival of the pandemic, low seeding rate, youthful demographics and possible prior immunity to coronavirus-like infections Njenga et al. (2020). Nevertheless, the pandemic is accelerating in Africa and passed 2.0 million confirmed cases on November 13, 2020 according to the Africa-CDC (2020) situation report. In Ethiopia, the first imported COVID-19 case, a Japanese person who travelled from Burkina Faso, was detected and reported by the Ministry of Health on March 13, 2020, Institute (2020). To control the spread of the disease, the Ethiopian government took immediate action, by closing schools, banning sporting events and mass gatherings, three days later. It took 79 days to reach the first 1000 cases, but 10,000, 20,000, 40,000 and 80,000 cases were attained in shorter periods of 51, 16, 18 and 44 days, respectively. This shows that the number of cases has been increasing over time, surpassing 100,000 by the mid of November, 2020. As the pandemic accelerates globally, many African countries, including Ethiopia, have been implementing different non-pharmaceutical interventions (NPIs) to contain the spread of the disease. The WHO is leading a global COVAX initiative for accelerated development of promising vaccine candidates that may be available in early 2021, and experimental therapies are also undergoing clinical trials for safety before licensing. However, at this point in time, the most effective means to control the spread of COVID-19 remains the implementation of different NPIs to break chains of transmission Fong et al. (2020b); Ryu et al. (2020); Lai et al. (2020); Flaxman et al. (2020); Ferguson et al. (2020). The transmission pathways of COVID-19 from person to person are: i) close contact through respiratory droplet, ii) direct contact with infected persons, and iii) contact with contaminated formites (objects and surfaces) Kassa et al. (2020). Public health measures are intended to diminish these transmission mechanisms.

Mathematical models have been previously used with success in understanding the transmission dynamics and control mechanisms of infectious diseases (Anderson and May (1979)). To understand the early transmission dynamics of COVID-19 under different scenarios, a number of mathematical models have been proposed in the literature, Prem et al. (2020); Li et al. (2020); Walker et al. (2020); Ivorra et al. (2020); Ngonghala et al. (2020); Nicholas et al. (2020). Those epidemiological and mathematical models contributed important insight for public health decision-makers to enforce different mitigation strategies in different countries. As COVID-19 has spread worldwide, most countries are utilising mathematical models to inform on public health measures. Many countries (for example the UK, China, Germany, USA, Morocco) revised their public health measures based on COVID-19 modelling results. We believe that, due to the difference in the age-structure of the population, social interaction and life style in Ethiopia, mathematical models developed in other countries may not work to study the dynamics of disease in lower income settings. This study employs the use of mathematical models to simulate the spread and interruption of transmission of the disease in Ethiopia.

Assessment of implemented and proposed intervention strategies for combating COVID-19, and predicting the number of new cases and deaths is a major challenge to both the public and scientific community. Banholzer et al. (2020) and Flaxman et al. (2020) estimated the impact of NPIs based on confirmed cases of COVID-19 in several countries. Similarly, Ferguson et al. (2020) studied the impact of NPIs to reduce COVID-19 mortality and healthcare demand. To our knowledge, there has been very limited study of the impact of NPIs on the disease dynamics in the local context of Ethiopia. The only published study, done by Siraj et al. (2020), focused on small, medium and large clusters of the population with social distancing face masking and contact tracing implementation at different proportion. This study did not quantify the impact of hand washing measures on transmission dynamics. Further, their study did not provide the estimated number of cases under different stages of the disease. Takele (2020) implements the Autoregressive Integrated Moving Average (ARIMA) modeling to project COVID-19 prevalence patterns in East Africa Countries, mainly Ethiopia, Djibouti, Sudan and Somalia. While, her projection model did not take into account the impact of NPIs with different adherence level on the predicted number of cases.

Our study proposes a modified mathematical model of the classic SEIR model (Anderson and May (1979)) and this modified model is used to compare the effect of different NPIs individually and in combination. The proposed model classifies the human population into eight non-overlapping stages of infection: Susceptible (*S*), Exposed (*E*), Asymptomatic infectious (*I*_*a*_), Symptomatic infectious (*I*_*s*_), Isolation of cases with mild/moderate health condition (at home or hospital) (*HI*_*m*_), Hospitalized with critical health condition (*H*_*c*_), Recovered from the disease (*R*), and Death due to COVID-19 (*D*).

Our proposed model (1) has a number of advantages: i) provide estimated time and size of the peak under the implementation of NPIs with different adherence level in urban and rural population settings, ii) proposed model differentiates asymptomatic and symptomatic infectious which influences the number of ICU cases and death due to the disease, iii) the effect of indirect transmission of the disease through contaminated environment is taken into account, and iv) provide the estimated impact of individual and synergy public health measures on the dynamics of the disease. The projected number of people in each stage of the disease at the peak period and the time of the peak in urban and rural areas of Ethiopia helps the government to choose and enforce better intervention mechanisms.

## 2 Materials and Methods

### 2.1 Representation of Proposed Mathematical Model

In our modeling framework, the population is divided into different compartments according to the infection status of individuals: susceptible (S), exposed (E), asymptomatic infected (*I*_*a*_), symptomatic infected (*I*_*s*_), isolated at home or hospital with moderate health condition (*IH*_*m*_), hospitalized with severe health condition (*H*_*c*_), recovered from the disease (R), and death (D) due to the disease. Contaminated environment is also a means of transmission for COVID-19 as the virus can stay up to several days on different surfaces van Doremalen et al. (2020). To account for the impact of contaminated objects in the transmission, an additional compartment for contaminated environment is included. Paramter description with these assumed initial values are presented in Table 1. Those values were extracted from a number of key papers and reports of the situation in Ethiopia (Institute (2020); Goshu et al. (2020); Baye (2020); Kebede et al. (2020); Siraj et al. (2020)) and epidemiological scientific facts of COVID-19.

By assuming that the total population is *N* = *S* + *E* + *I*_*a*_ + *I*_*s*_ + *IH*_*m*_ + *H*_*C*_ + *R* + *D*, the corresponding system of differential equations of our proposed model is given by

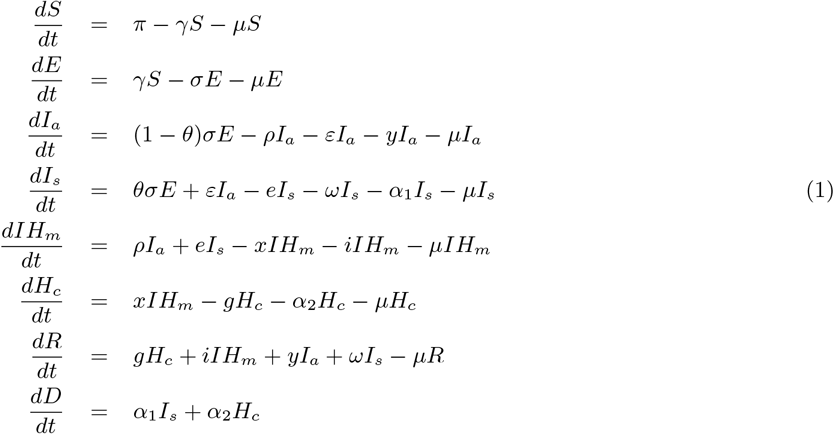

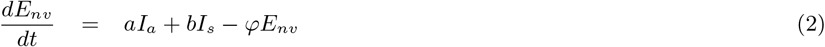

where the force of infection, *γ*, is obtained using the following formula:

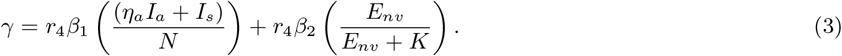

**Table 1:** Description and values of initial model parameters 1

| Parameter | Description | Value |  |  |
| --- | --- | --- | --- | --- |
|  |  | Global-level | Ethiopia |  |
| $\gamma$ | force of infection | | estimated | |
| $\beta_1$ | rate of disease transmission directly from human | .807 | | |
| $\beta_2$ | rate of disease transmission from<br>contaminated environment | 0.4 | | |
| $\eta_a$ | relative infectiousness of person in $I_a$ | 0.5 | | |
| $\epsilon$ | proportion of the exposed that goes to $I_a$ | 0.2 | | |
| $\theta$ | proportion of the exposed transfer to $I_s$ | 0.8 | | |
| $\sigma$ | rate at which exposed individuals are detected and move to $I$ | 0.2 | | |
| $\alpha_2$ | Disease induced death rate | 0.15 | 0.015 | |
| a | Shedding rate from the asymptomatic Infectious class | 1/8 |  | Ferguson et al. |
| b | Shedding rate from the symptomatic infectious class | 1/10 |  |  |
| $\varphi$ | Virus decays from the environment | 1/4 | | |
| $\varepsilon$ | rate of $I_a$ develop symp. and going to $I_s$ | 0.4 | | |
| e | rate of detected from $I_s$ going to $IH_m$ | 0,8 | | |
| $\rho$ | rate of detection and isolating or go to $IH_m$ | 0.4 | | |
| y | Rate of recovery from $I_a$ | 0.2 | | |
| $\omega$ | Rate of recovery from $I_s$ | 0.4 | | |
| x | Rate of transfer from $IH_m$ to $H_c$ | 0.08 | 0.01 | |
| $\alpha_1$ | Death Rate of transfer from $I_s$ | 0.1 | | |
| g | Rate of Recovery after getting critical | 0.02 | 0.08 |  |
| i | Rate of Recovery after getting mild | 0.4 | 0.6 |  |
| $\mu$ | Natural death rate | 0.0076 | 0.0064 | Organ |
| $\pi$ | Natural birth rate | 0.018 | 0.03 | |

In Equation (3, *β*_1_ and *β*_2_ are effective contact rates leading to COVID-19, *η*_*a*_ is relative infectiousness per contact for asymptomatic patients, *r*_4_ = (1−*cFM*) *(1−*SD*)*(1−*HW*), where *SD* represents the proportion of the population who practice physical distancing, *HW* represents the proportion of the population who implements hand washing, *FM* is coverage of wearing masks, *c* is efficacy of face mask wearing at reducing transmission, and *K* is the virus concentration in the environment that yields 50% of chance for a susceptible individual to catch the viral infection from that source, Kassa et al. (2020). The model accounts for a distinction between non-diagnosed individuals *I*_*a*_ and *I*_*s*_, who can readily spread the infection because they are not in isolation, and hospitalized individuals *HI*_*m*_ and *H*_*c*_, who transmit the disease less thanks to isolation and complying with strict rules, either in hospital or at home.

### 2.2 Well-posedness of the System and sensitivity analysis

To check the mathematical and biological meaningfulness of the model, both positivity and boundedness properties of the proposed model are assessed as follows. Let the domain space be defined by

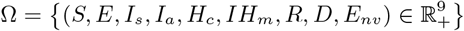

#### Theorem 2.1.

*int*(Ω) = *{*(*S, E, I*_*s*_, *I*_*a*_, *H*_*c*_, *IH*_*m*_, *R, D, E*_*nv*_) |*S* > 0, *E* ≥ 0, *I*_*s*_ ≥ 0, *I*_*a*_ ≥ 0, *H*_*c*_ ≥ 0, *IH*_*m*_ ≥ 0, *R* ≥ 0, *D* ≥ 0, *E*_*nv*_ ≥ 0)*} is the positive invariant set of the system* (2).

The proof of theorem 2.1 is presented in the Appendix and the theorem implies well-posedness of the system.

#### Sensitivity analysis of model parameters

In order to identify parameters which significantly influence the model system, sensitivity analysis was performed. The uncertainty and sensitivity analysis is done by using Partial Rank Correlation Coefficient (PRCC) analysis with N=10,000 samples for various input parameters Wu et al. (2013); Marino et al. (2008). The partial derivative of the threshold value *R*_*o*_ with respect to the input parameters were computed by varying the parameters around normal values.

### 2.3 System Equilibria and Stability of the System

#### 2.3.1 Disease Free Equilibrium

Equilibrium points are those points where each equation in the system in Equation (1) is equal to zero. Equating the right hand side of each equation in Equation (1) to zero will provide disease-free and endemic equilibrium points. In this subsection, the disease-free equilibrium point is described as follows.

When there is no COVID-19 in the population, the disease-free equilibrium point is given by

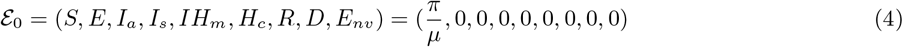

In this study, to compute the basic reproduction number, Diekmann et al. (1990), next-generation method for the disease-free equilibrium is employed. The basic reproduction number is the average number of secondary infection cases produced in a completely susceptible population by a typical infectious individual. According to the definitions stated in Diekmann et al. (1990) and van den Driessche and Watmough (2002a), in the next-generation method, *R*_0_ is the spectral radius of the next-generation matrix which is given by

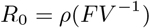

where *F* is the Jacobian of the rate of appearance of new infections in the infected compartment, denoted by *f* and *V* is the Jacobian of *v* = *v*^−^ − *v*^+^ that represents all infection transfer interactions into (*v*^+^) and out (*v*^−^) of these compartments.

The infection compartments of our model are (*E, I*_*a*_, *I*_*s*_, *IH*_*m*_, *H*_*c*_, *E*_*nv*_)

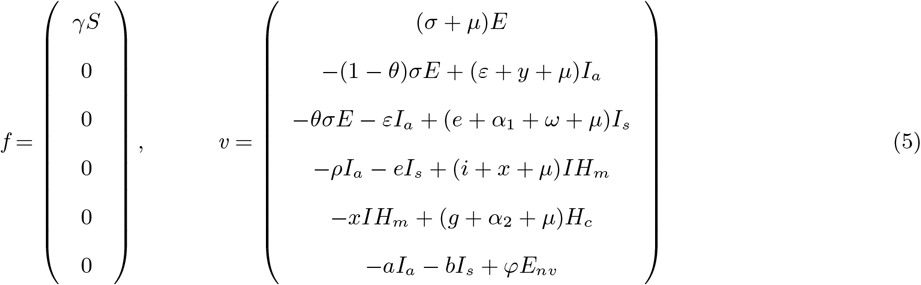

Computing the Jacobian of *f* and evaluating it at the disease-free equilibrium point using the force of infection, we have,

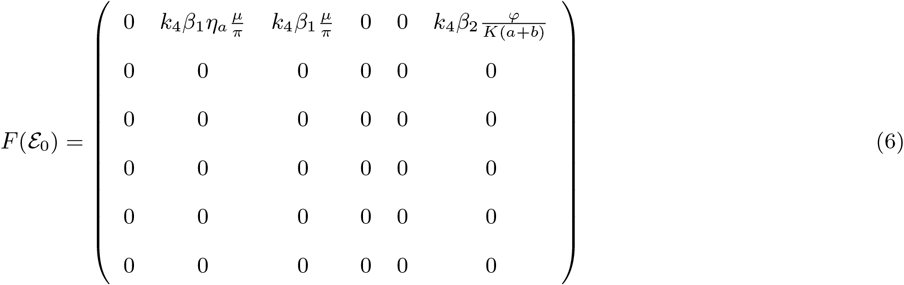

and the Jacobian of *v* evaluated at the disease free equilibrium point using the force of infection is given by the matrix

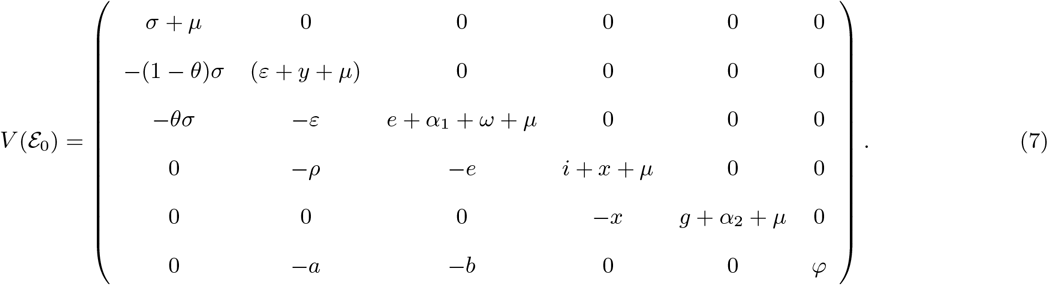

Finding the inverse of *V* and computing the product *FV* ^−1^, the next generation matrix is given by

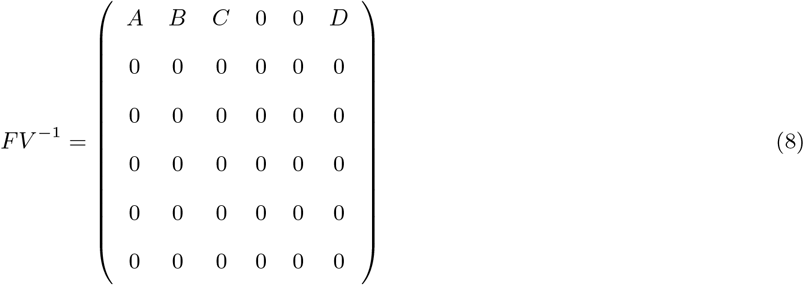

where for

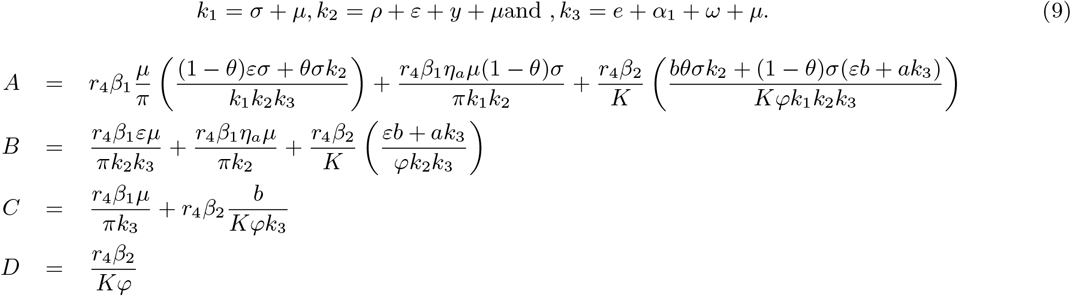

The corresponding Spectral radius of the matrix *FV* ^−1^ is given by *R*_0_ = *ρ*(*FV* ^−1^) = *A*, where

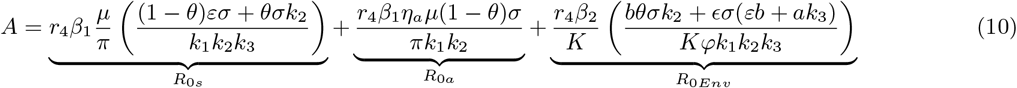

which can be expressed as *A* = *R*_0_ = *R*_0*a*_ + *R*_0*s*_ + *R*_0*Env*_ and the quantity *R*_0_ is the basic reproduction number of the Model (2). The quantity *R*_0_ is the sum of the constituent reproduction numbers associated with the number of new COVID-19 cases generated by symptomatically infectious humans (*R*_0*s*_), asymptomatically infectious humans (*R*_0*a*_) and contaminated environment (*R*_0*E*_*nv*).

##### Theorem 2.2.

*The disease-free equilibrium (DFE) of model 2 is locally asymptotically stable if R*_0_ *<* 1, *and unstable if R*_0_ > 1.

*Proof*. It follows from Theorem 2 of van den Driessche and Watmough (2002b)

The implication of Theorem 2.2 is that a small introduction of COVID-19 cases will not generate a COVID-19 outbreak if the basic reproduction number (*R*_0_) is less than unity.

##### Theorem 2.3.

*The disease-free equilibrium of model 2 is globally asymptotically stable if R*_0_ *≤* 1.

#### 2.3.2 Endemic Equilibrium

The endemic equilibrium is attained in the presence of the disease, which is denoted by:

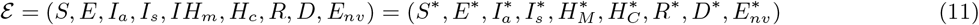

and the values of 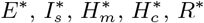 and 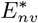 can be computed from the equations:

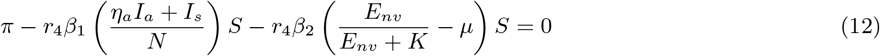

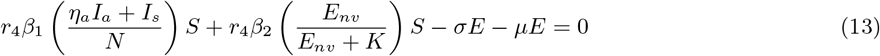

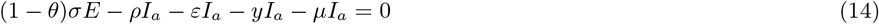

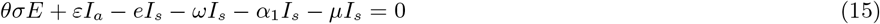

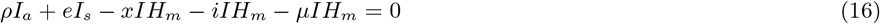

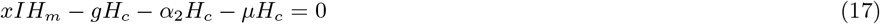

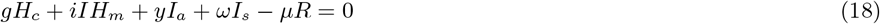

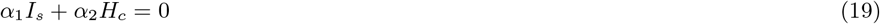

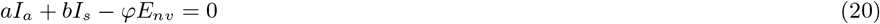

From Equation (14), solving for *E*\*, we have

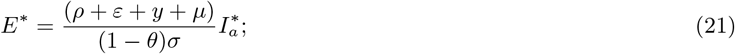

from Equation (15) solving for 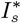 we have

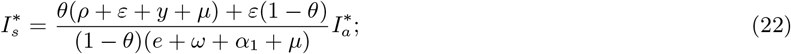

from Equation (16), solving for 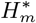 we have

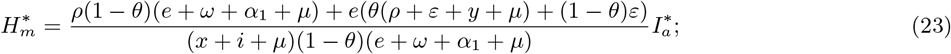

solving for 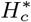 from Equation (17) we have

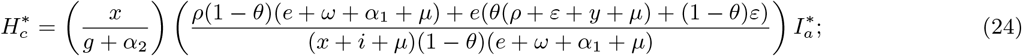

solving Equation (18) for *R*\* we have

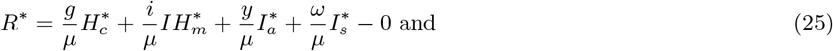

by solving Equation (20) for 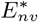 we can get

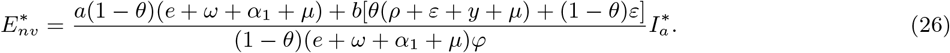

### 2.4 Projection of Cases using Proposed Model

The proposed modified SEIR model described in section 2.1 is used to project the number of COVID-19 related cases in Ethiopia. Projecting the number of active COVID-19 cases under the implementation of different NPIs with different adherence level is important for policy makers to be able to mitigate against the disease. Projecting the number of asymptomatic and symptomatic people is important, as these two numbers have huge roles in the spread of the disease. Further, knowing the expected number of people who may require treatment under intensive care (ICU cases) will greatly help to guide policy makers in preparation of manpower and medical facilities such as ventilators. Due to the very limited number of available ventilators in Ethiopia (below 1000 ventilators in total), knowing the projected number of individuals who need ICU under the implementation of different NPIs with varying adherence level of hygiene, physical distancing and mask wearing are important to save lives. Many studies showed that most of the critically ill patients with COVID-19 are of older age and have more co-morbidities than the non-critically ill patients. Hence, projection is done for all active case, symptomatic cases, asymptomatic cases, ICU cases, and death due to the disease using the proposed model.

In our model, common COVID-19 related parameters were obtained from previous studies (Table 1). ^2^ Further, based on the 2016 Ethiopian demographic and health survey, 27.4% and 7.8% of the urban and rural population wash hands by soap, respectively. Thus, in the simulation, we assumed improved percentages of hygiene due to the awareness created by COVID-19 in both population settings. Due to differences in access to sanitation materials, lifestyle, cultural norms and other factors, adherence levels to the recommended NPIs are also different in the urban and rural populations. As a result, projection on the number of COVID-19 related cases is done separately for urban and rural populations of Ethiopia.

### 2.5 Data

In this study, data on daily number of COVID-19 cases, cumulative number of deaths, and number of critical patients was extracted from Ethiopian Public Health Institute website (www.ephi.gov.et/) and Ministry of Health official Twitter page(https://twitter.com/FMoHealth/) on daily basis. We used initial parameter (1)values describing the natural and clinical course of infection from published sources WHO (2020); Ngonghala et al. (2020); Ferguson et al. (2020); Kassa et al. (2020). The projection model considers daily number of cases in Ethiopia up to 210 days (October 08, 2020) since the first case of the disease was recorded.

### 2.6 Considered Non-Pharmaceutical Interventions (NPIs)

Based on the current evidence about COVID-19, implementing different NPIs are essential requirements for limiting the spread of the disease. The main objective of NPIs is to reduce the rate of transmission, thereby minimizing the size of the epidemic peak and delaying peak time, buying time for preparations in the healthcare system, and enabling the potential for vaccines and drugs to be developed, approved and used, Fong et al. (2020a); Ryu et al. (2020); Xiao et al. (2020); Flaxman et al. (2020); Lai et al. (2020). The NPIs considered in this study are physical-distancing, wearing face-masks and hygiene measures (hand washing)(see Table 2). We assume that implementing NPIs alone or in combination affects the rate of contact between uninfected people and infected people or objects. As a result, transmission probability of the virus from infected individuals or objects to the susceptible population is reduced.

**Table 2:** Summary of considered intervention mechanisms and their effect on reducing the spread of COVID-19 based on existing literature

| Intervention | Description | Reduction | References |
| --- | --- | --- | --- |
| Physical distancing | Creating ways to increase distance between people in settings where people commonly come into close contact with one another (i.e. schools, workplaces, events, spiritual centers, transports, etc). | 35 times | Courtemanche et al. (2020) |
| Face-masking | Wearing face-masks protects susceptible individuals from acquisition of infection. If infected individuals wear face-masks, it reduces their ability to transmit the virus. | 30-70% | Howarda et al. (2020),Ngonghala et al. (2020) |
| Hygiene (hand washing) | Frequent, thorough hand washing with soap and water is effective at preventing the spread of lipid-enveloped viruses, such as SARS-CoV-2. | 40% | Bloomfield et al. (2007),Health (2020) |

We evaluated the impact of the three NPIs, alone or in combination, on the time and size of the peak by varying the adherence level to each of them.

## 3 Results: Projection of number of COVID-19 Cases in Ethiopia

### 3.1 Projection of All Active Cases

By fixing the adherence levels to hygiene to 30% and 40% of the urban population based on the aforementioned reasons, the projected number of active COVID-19 cases was obtained by varying the percentages of social distancing and face mask coverage. Figure 2 presents the projected number of active COVID-19 cases with varying adherence levels of NPIs.

**Figure 1:**
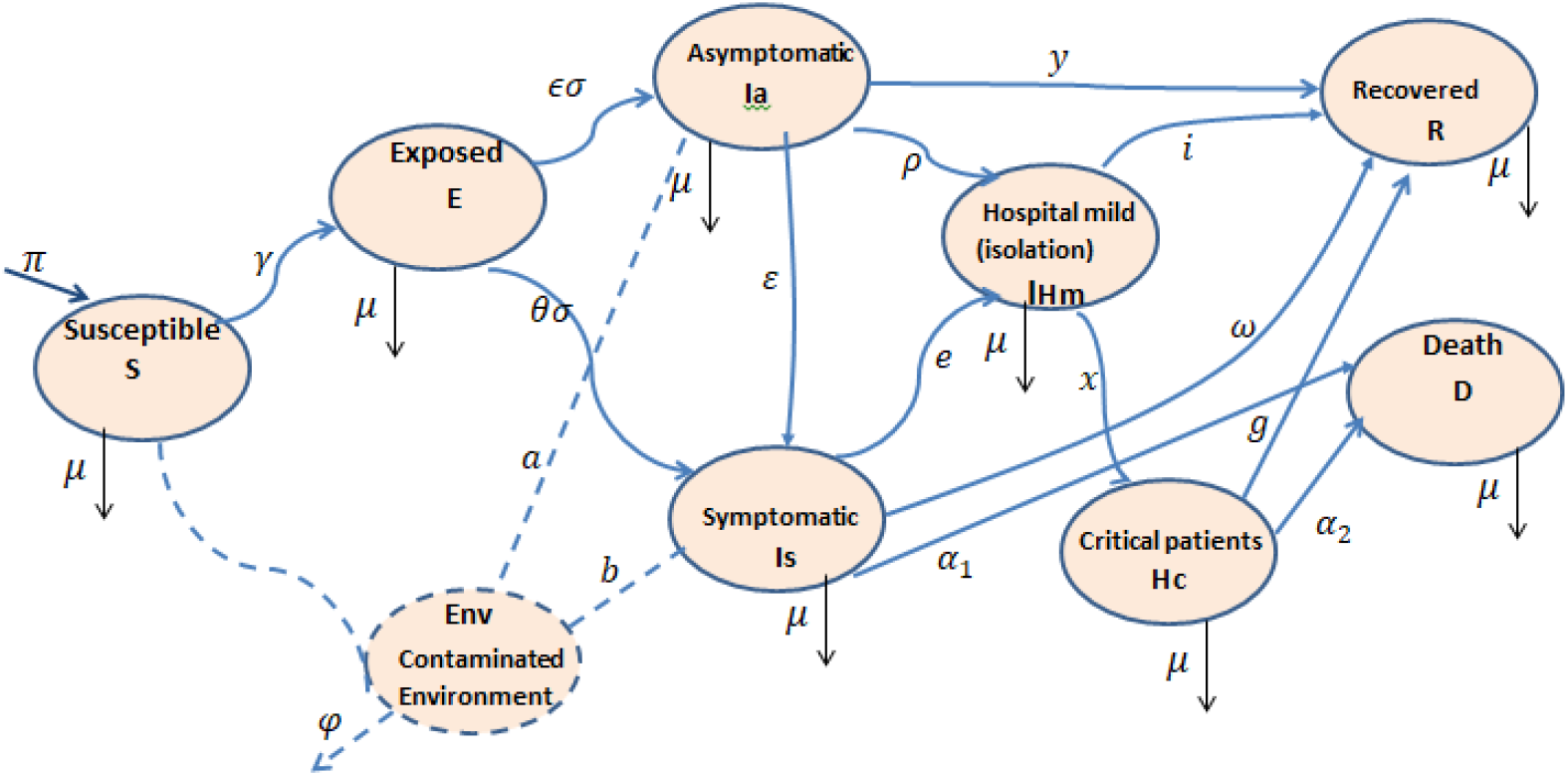
Flow diagram of the mathematical model showing the transition of individuals in different compartments based on infectious status

**Figure 2:**
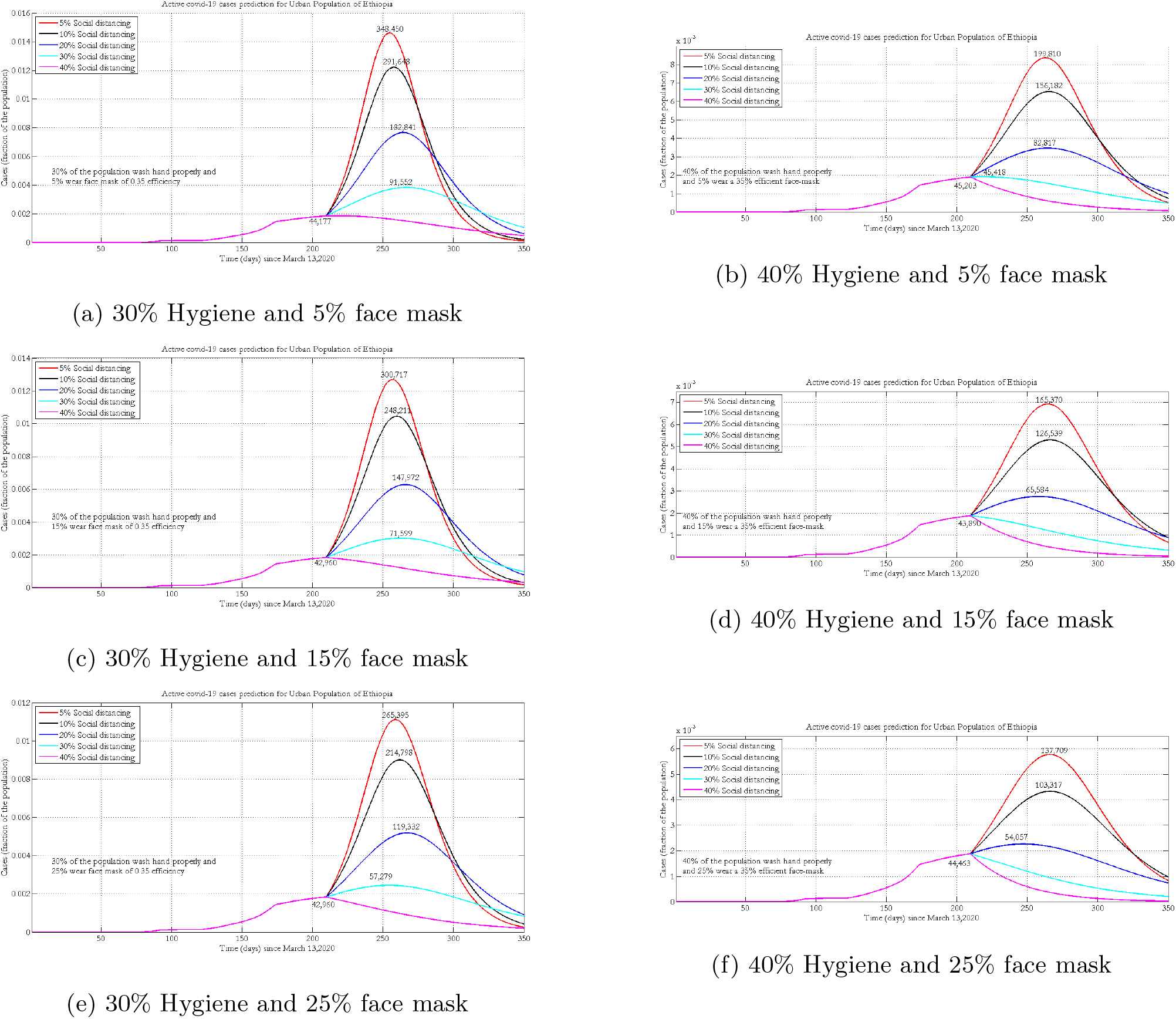
Projection of active COVID-19 cases in urban population of Ethiopia when only 5% (Figure 2a and 2b), 15% (Figure 2c 2d) and 25% (Figure 2e, 2f) of the population wear face masks with 35% efficacy

At 15% face mask coverage and with 10% of the population implementing social distancing measures, a 10% increase in hygiene will decrease the number of cases by 200,000 and delay the peak time. For a given implementation of wearing a face mask and hygiene, improving the percentage of social distancing by 10% will reduce the number of new COVID-19 cases by one-fifth. Increasing the number of people who wear face mask a by 10% will save on average 25,000 people from being infected by the disease (Figure 2).

In the rural parts of Ethiopia, physical distancing is a custom and part of the life style. This custom greatly minimizes the spread of the disease that is caused by contact and contaminated environment. Furthermore, due to low coverage of road accessibility (accessibility index=22%), in our simulation we assumed 20% of the population are protected until the end of the pandemic World-Bank (2016). Further, based on 2016 EDHS, 7.8% of the rural population wash their hands using soap. Figure 3 depicts the projected number of active cases under the implementations of varying physical distancing measures and wearing of face masks. In rural settings, people use home-made masks, and we assumed the efficacy of the masks is 0.2.

**Figure 3:**
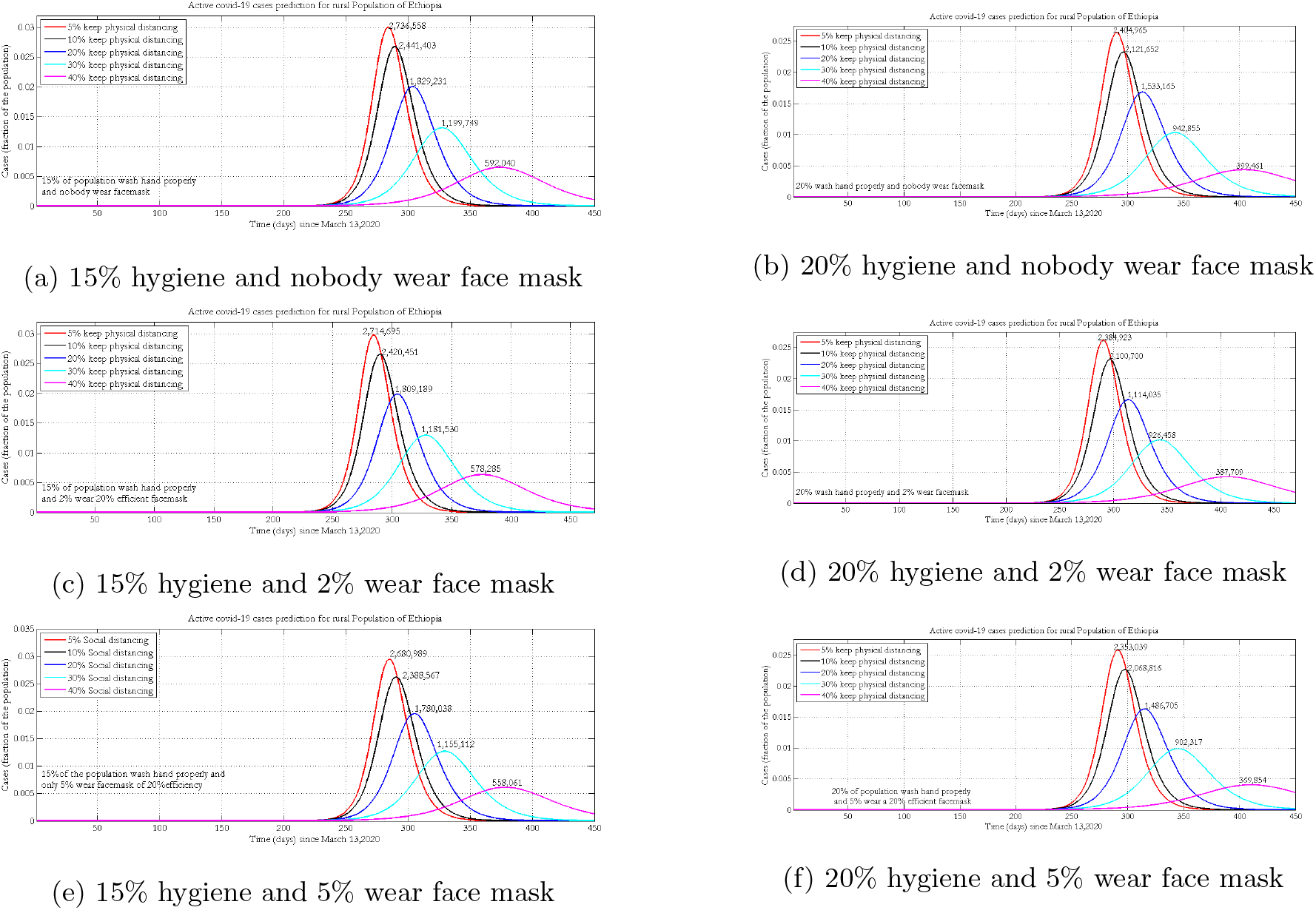
Projection of active COVID-19 cases in rural population of Ethiopia with 0% (Figure 3a,Figure 3b) 2% (Figure 3c,Figure 3d) 5% (Figure 3e,Figure 3f) wear face masks with 20% efficiency.

Keeping adherence to face mask wearing and physical distancing constant, a 5% increase in hygiene will decrease the number of cases by 200, 000 and shift the peak time to the future nearly by one month (Figure 3). For a given implementation of face mask wearing and hygiene, improving the percentage of physical distancing by 10% will reduce the number of new COVID-19 cases by one-fourth.

### 3.2 Projection of Symptomatic and Asymptomatic Cases

The following projected number of symptomatic (broken line) and asymptomatic (solid line) cases were simulated by assuming 30% and 40% of the urban population wash hands with soap, with different face mask coverage (5%, 15%and 25%) with 35% mask efficacy.

When 40% of the urban population wash hands with soap or keep hands clean using sanitizers, 25% wear face masks, improving the implementation of physical distancing greatly reduces both symptomatic and asymptomatic infections. By increasing the percentage of physical distancing in the population from 10% to 20%, the peak size will decrease by half for both infection compartments (Figure 4).

**Figure 4:**
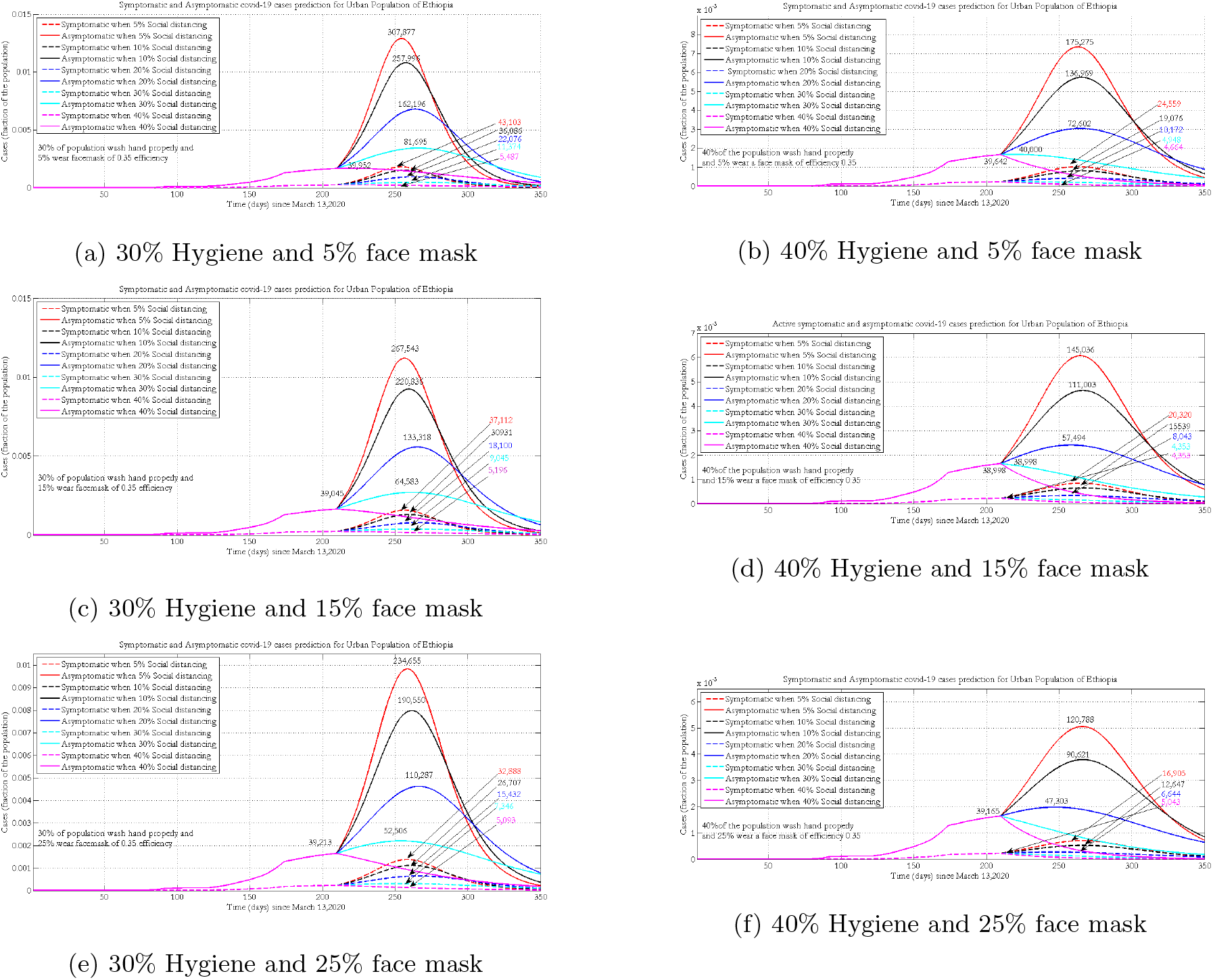
Projection of symptomatic and asymptomatic infectious cases in urban populations of Ethiopia when 5% (4a 4b) 15% (4c 4d) and 25% (4e,4f) of them wear face masks with 35% efficacy.

For a given coverage of face mask wearing and implementing physical distancing, improving hygiene by 10%, highly reduces the number of active cases and delays the peak time of the pandemic. For both hygiene scenarios, for a given level of face mask wearing, increasing adherence to physical distancing profoundly reduces the number of symptomatic and asymptomatic cases. A maximum number of asymptomatic infections will be observed if there is poor implementation of physical distancing and face mask wearing. Regardless of other NPIs, COVID-19 will be suppressed if 40% of the population implements physical distancing properly.

Figure 5 presents the projected number of symptomatic (broken line) and asymptomatic (solid line) cases in rural Ethiopia by varying the level of face mask wearing and physical distancing while fixing hygiene practice in the community at 15% and 20%. Increasing proper hand washing behaviour from 15% to 20% could shift the peak by around 2 months. Implementing physical distancing, hygiene, and face mask wearing at 40%, 20% and 5% levels, respectively, helps to control the pandemic under the Ethiopian health system capacity and shifts the peak forward to 2021. At a given level of implementation of hand washing and wearing of face masks, increasing the physical distancing by 10% could shift the peak time by around 40 days (Figure 5).

**Figure 5:**
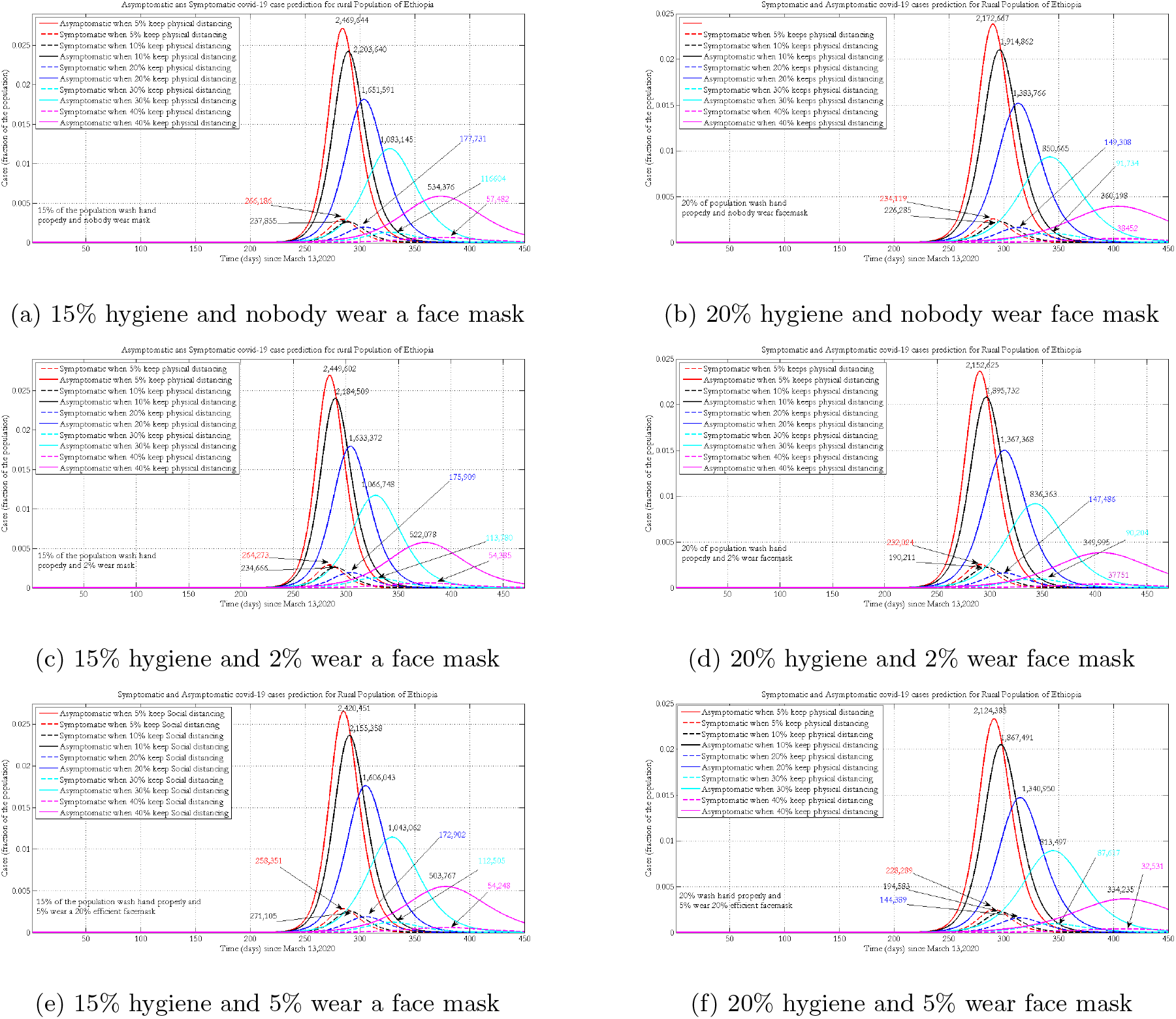
Projection of symptomatic and asymptomatic infectious cases in rural population of Ethiopia when 0%(Figure 5a, Figure 5b) 2% (Figure 5c, Figure 5d) 5%(Figure 5e,Figure 5f) of the rural population properly wear a face mask

### 3.3 Projection of Critical Cases

Figure 6 presents the projected number of ICU cases by keeping the hygiene coverage constant (at 30%and 40%) and varying adherence to physical distancing and wearing of face masks at different levels in urban population of Ethiopia.

**Figure 6:**
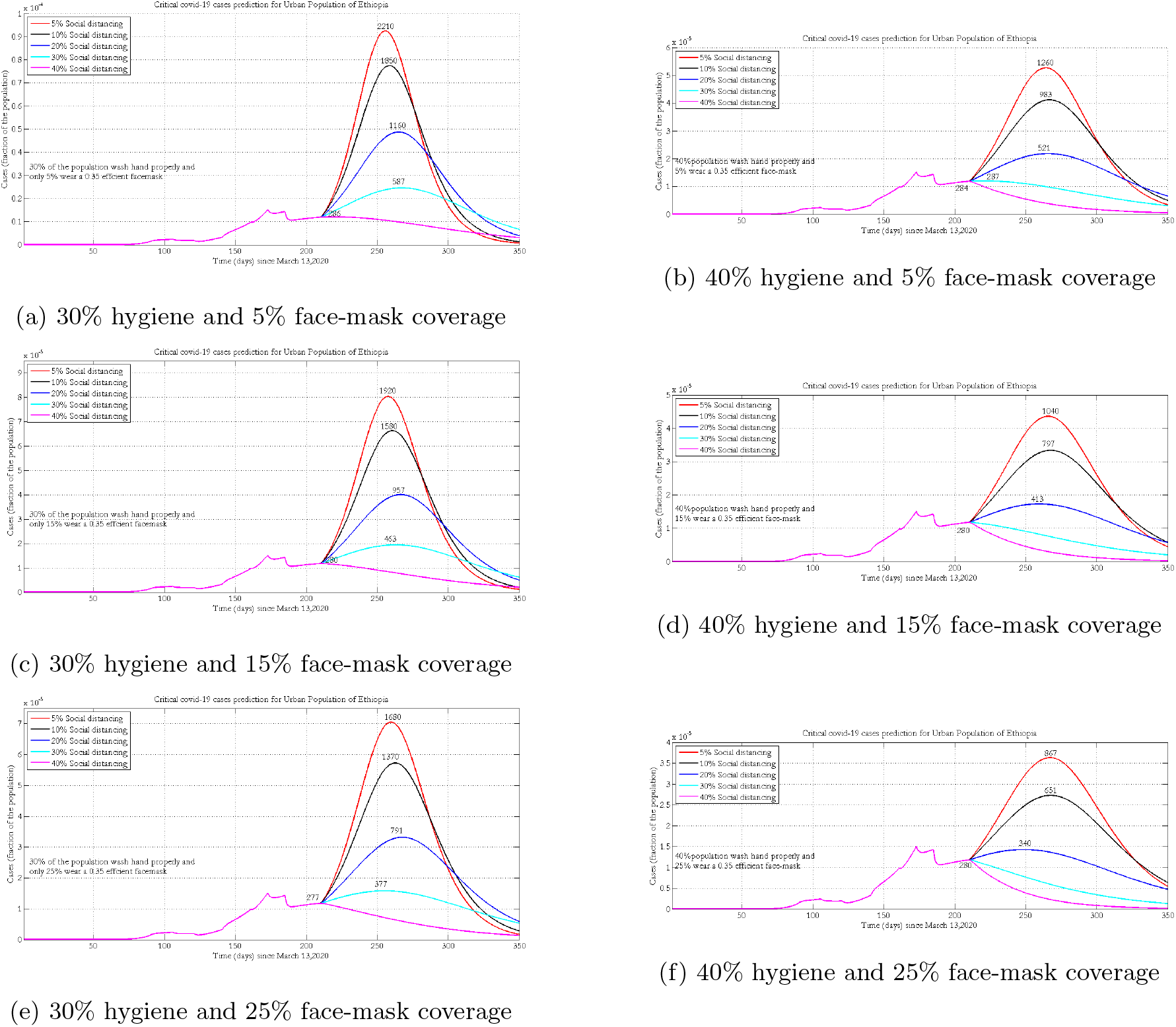
Projection number of urban population who will need intensive care when 5%(Figure 6a Figure 6b) 15% (Figure 6c,Figure 6d) and 25% (Figure 6e, Figure 6f) of them wear face mask

If 15% of the population wears masks and 30% implement physical distancing, improving hygiene by 10% could reduce the number of ICU cases from 463 to 280 (Figure 6). Proper implementation of wearing face masks and hand washing could flatten the curve. However, to bring critical cases under the Ethiopian health care capacity, people should also practice social distancing.

It is not too late to suppress the disease if strict implementation of NPIs is practiced (40% hand wash, 40% physical distancing and 25% wearing masks) in all urban populations of Ethiopia.

In the rural population the projected number of critical cases could exceed the available number of ventilators in our health system (Figure 7). To contain critical cases, people should strictly implement the three NPIs or the government should enforce additional public health measures. Since the time of the peak in rural populations is different from that of urban populations, the health system can make preparations for the transport system to bring these critical cases to heath facilities in urban centers. The two extreme scenarios show that improving hygiene by 5% and physical distancing by 40% could reduce the total?at peak time? number of critical cases from 2,361 to 1,569 (assuming 5% wear masks). Strict implementation of the three NPIs will shift the peak time forward by three months (Figure 7).

**Figure 7:**
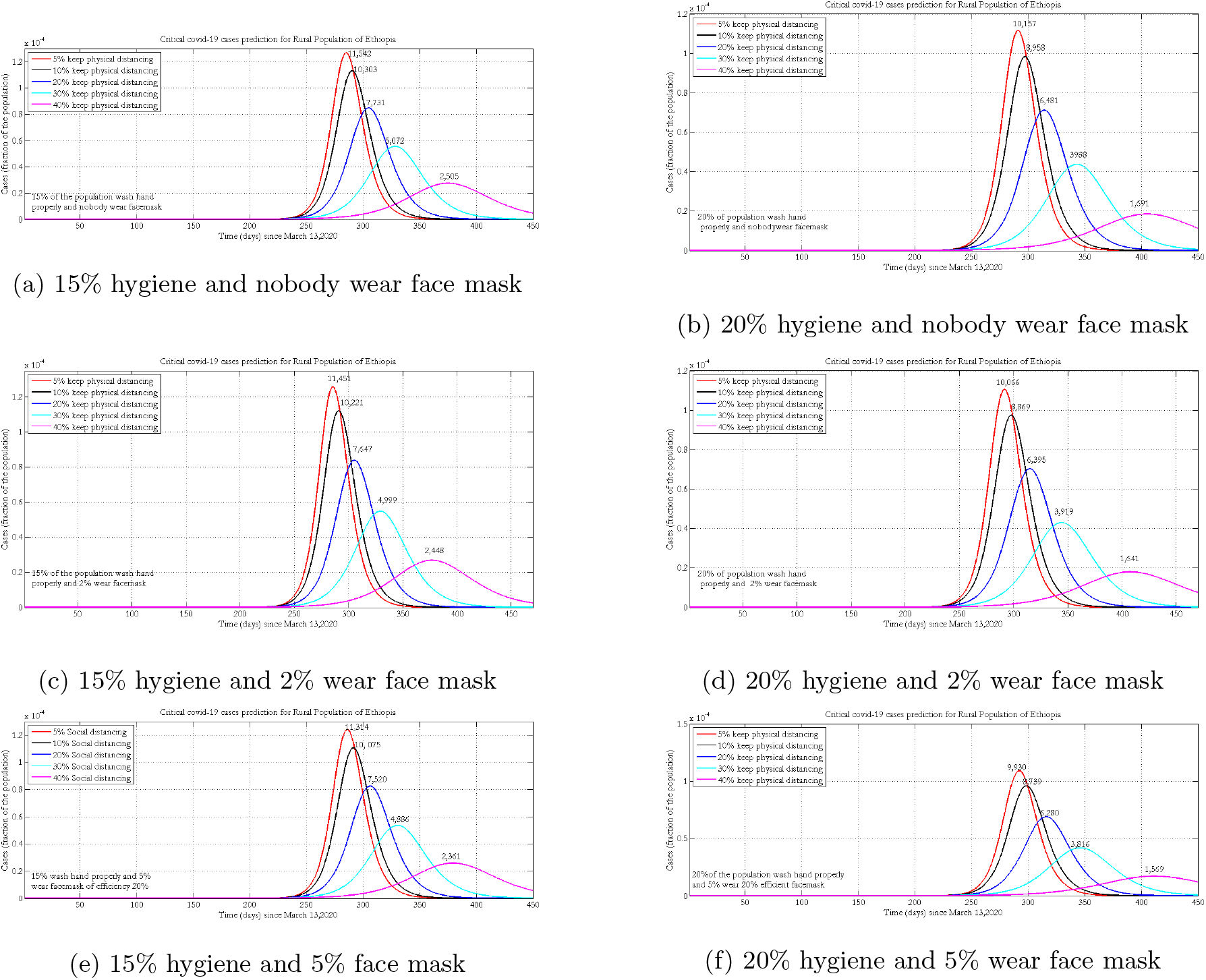
Projection of intence care or critical cases in the rural population of Ethiopia when 0% (Figure 7a, Figure 7b) 2% (Figure 7c, Figure 7d) 5%(Figure 7e,Figure 7f) of the population are wearing facemask with 20% efficacy.

### 3.4 Projection of Cumulative Death Due to COVID-19

Figure 8 presents the projected number of deaths due to COVID-19 under different scenarios in urban populations of Ethiopia.

**Figure 8:**
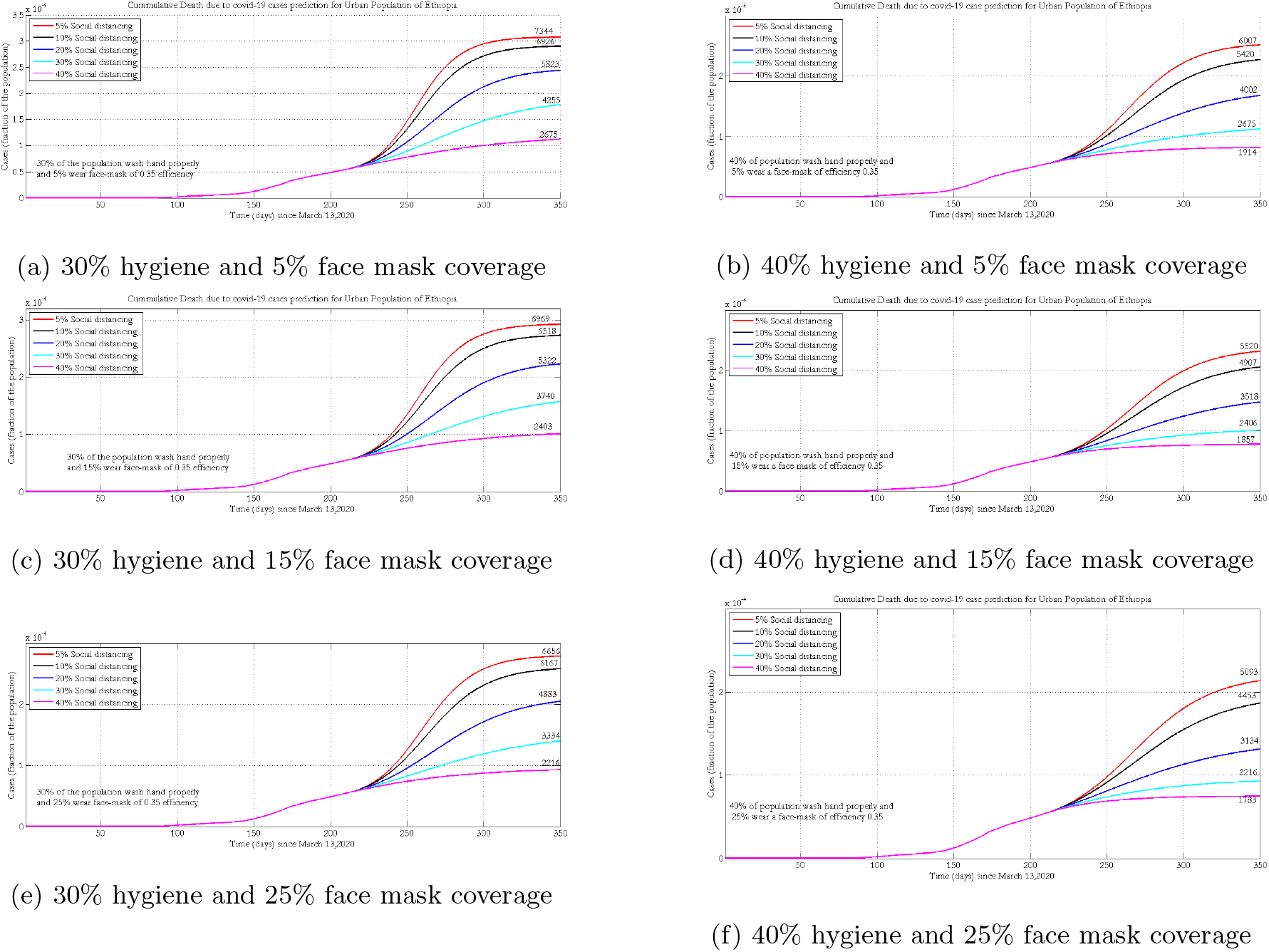
Projection of deaths under the implementation of different NPIs when 5%(Figure 8a, Figure 8b) 15% (Figure 8c,Figure 8d) and 25% (Figure 8e, Figure 8f)of them wear face masks properly.

The number of deaths due to COVID-19 could be reduced if more than 40% of the populations practice physical distancing. The projected number of cumulative deaths under the implementation of different adherence levels of physical distancing and face mask wearing at fixed (15% and 20%) levels of hygiene are presented in Figure 9.

**Figure 9:**
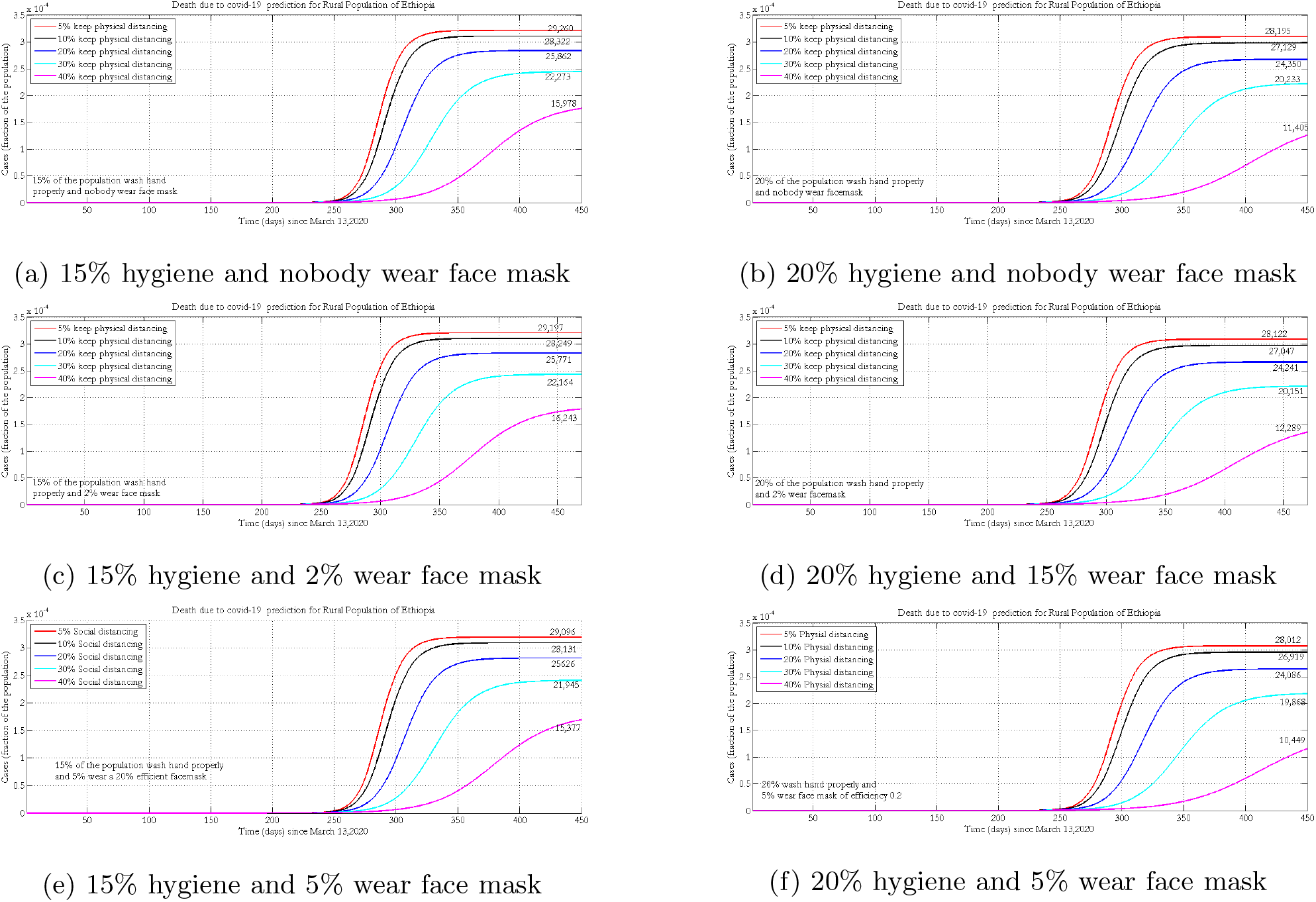
Projected number of cumulative death due to covid-19 in rural Ethiopia when nobody (Figure 9a,Figure 9b) 2% (Figure 9c, Figure 9d) 5%(Figure 9e,Figure 9f) of the population wear face mask of 0.2 efficiency.

Practicing recommended NPIs properly (40% physical distancing 5% face mask and 20% proper hand washing) could greatly reduce the number of death due to COVID-19. Table 3 and Table 4 presents summary of the projected number of active COVID-19 and ICU cases under different adherence levels of hygiene, wearing-masks and physical-distancing in the urban and rural population of Ethiopia, respectively.

**Table 3:** Summary of predicted number of active Covid-19 and ICU cases (in thousands) under different adherence levels of wearing-masks and physical-distancing in the Urban population of Ethiopia.

| Distancing | Percentage of population who wear face masks of efficiency 35%, Hygiene=30% |  |  |  |  |  |  |  |  |  |  |  |
| --- | --- | --- | --- | --- | --- | --- | --- | --- | --- | --- | --- | --- |
|  | Projected Number of all active Cases |  |  |  |  |  | Projected number of active ICU cases |  |  |  |  |  |
|  | 5 |  | 15 |  | 25 |  | 5 |  | 15 |  | 25 |  |
|  | Size | Period | Size | Period | Size | Period | Size | Period | Size | Period | Size | Period |
| 5 | 348 | Nov. | 300 | Nov. | 265 | Nov. | 2.21 | Nov. | 1.92 | Nov. | 1.68 | Nov. |
| 10 | 292 | Nov. | 248 | Nov. | 214 | Nov. | 1.85 | Nov. | 1.58 | Nov. | 1.37 | Dec. |
| 20 | 183 | Dec. | 147 | Dec. | 119 | Dec. | 1.16 | Dec. | 0.95 | Dec. | 0.79 | Dec. |
| 30 | 91 | Dec. | 71 | Dec. | 57 | Nov. | 0.5 | Dec. | 0.46 | Dec. | 0.37 | Nov. |
| 40 | 44 | Oct. | 42 | Oct. | 42 | Oct. | 0.2 | Oct. | 0.2 | Oct. | 0.27 | Nov. |
|  | Percentage of population who wear-facemasks of efficiency 35%, Hygiene=40% |  |  |  |  |  |  |  |  |  |  |  |
| 5 | 199 | Dec. | 165 | Dec. | 137 | Dec. | 1.26 | Dec. | 1.04 | Dec. | 0.8 | Dec. |
| 10 | 156 | Dec. | 126 | Dec. | 103 | Dec. | 0.9 | Dec. | 0.7 | Dec. | 0.6 | Dec. |
| 20 | 82 | Dec. | 65 | Nov. | 57 | Nov. | 0.5 | Dec. | 0.4 | Nov. | 0.3 | Nov. |
| 30 | 45.4 | Oct. | 44 | Oct. | 44 | Oct. | 0.2 | Oct. | 0.2 | Oct. | 0.2 | Oct. |
| 40 | 45.2 | Oct. | 44 | Oct. | 44 | oct. | 0.2 | Oct. | 0.2 | Oct. | 0.2 | Oct. |

**Table 4:** Summary of predicted number of active Covid-19 and ICU cases (in thousands) under different adherence levels of wearing-masks and social-distancing in the rural population of Ethiopia

| Face-masking | Percentage of population who wear-facemasks of efficiency 20%, Hygiene=15% |  |  |  |  |  |  |  |  |  |  |  |
| --- | --- | --- | --- | --- | --- | --- | --- | --- | --- | --- | --- | --- |
|  | Projected Number of all active Cases |  |  |  |  |  | Projected number of active ICU cases |  |  |  |  |  |
|  | 0 |  | 2 |  | 5 |  | 0 |  | 2 |  | 5 |  |
|  | Size | Period | Size | Period | Size | Period | Size | Period | Size | Period | Size | Period |
| 5 | 2736 | Dec. | 2714 | Dec. | 2680 | Dec. | 11 | Dec | 11 | Dec | 11 | Dec |
| 10 | 2441 | Dec. | 2420 | Dec. | 2388 | Dec | 10 | Dec. | 10 | Dec. | 10 | Dec. |
| 20 | 1829 | Jan 21 | 1809 | Jan 21 | 1780 | Jan. 21 | 7 | Jan 21 | 7 | Jan 21 | 7 | Jan 21 |
| 30 | 1199 | Feb. 21 | 1181 | Feb.21 | 1155 | Feb. 21 | 5 | Feb 21 | 5 | Feb 21 | 5 | Feb.21 |
| 40 | 592 | Mar 21 | 578 | Mar. 21 | 558 | Mar. 21 | 2 | Mar 21 | 2 | Mar 21 | 2 | Mar 21 |
|  | Hygiene improved from 15% to 20% |  |  |  |  |  |  |  |  |  |  |  |
| 5 | 2404 | Dec. | 2384 | Dec. | 2353 | Dec. | 10 | Dec. | 10 | Dec. | 9 | Dec. |
| 10 | 2121 | Jan 21 | 2100 | Jan 21 | 2069 | Jan 21 | 8 | Jan 21 | 8 | Jan 21 | 8 | Jan 21 |
| 20 | 1533 | Jan 21 | 1514 | Jan 21 | 1485 | Jan 21 | 6 | Jan 21 | 6 | Jan 21 | 6 | Jan 21 |
| 30 | 942 | Feb 21 | 926 | Feb 21 | 902 | Feb 21 | 3 | Feb 21 | 3 | Feb 21 | 3 | Feb 21 |
| 40 | 399 | Mar 21 | 387 | Mar 21 | 370 | Mar 21 | 1 | Mar 21 | 1 | Mar 21 | 1 | Mar 21 |

### 3.5 The Basic Reproductive Number

The basic reproductive number (*R*_*o*_) measures the degree of spread of SARS-CoV-2. If this number is greater than one, the disease will spread out if additional public health measures are not taken.

Table 5 and 6 presents estimated *R*_*o*_ at different time points by taking into account enforced NPIs since the onset of the disease in urban and rural Ethiopia, respectively. This number was higher in the first and 10^*th*^ week of the pandemic in urban Ethiopia. Further, compared with the rural population, at the beginning of the pandemic *R*_*o*_ was higher in the urban areas of Ethiopia.

**Table 5:** Estimated *R*_*o*_ at different time points since the onset of COVID-19 in urban population of Ethiopia

| Days | 2-60days | 61-90days | 91-105 days | 106-120days | 121-140days | 141-160days | 161-172days |
| --- | --- | --- | --- | --- | --- | --- | --- |
| $R_o$ | 0.605 | 0.95 | 0.46 | 0.409 | 0.58 | 0.53 | 0.568 |
| Days | 173-184days | 185-210days |  |  |  |  |  |
| $R_o$ | 0.45 | 0.45 | | | | | |

**Table 6:** Estimated *R*_*o*_ at different time points since the onset of COVID-19 in rural population of Ethiopia

| days | 2-60days | 61-90 days | 91-105 days | 106-120days | 121-140 days | 141-160 days |
| --- | --- | --- | --- | --- | --- | --- |
| $R_o$ | 0.61 | 0.5593 | 0.4645 | 0.41 | 0.58 | 0.54 |
| Days | 161-172 days | 173-180 days | 181-215 days |  |  |  |
| $R_o$ | 0.53 | 0.40 | 0.43 | | | |

Table 7 presents summary of *R*_*o*_ under the implementation of different NPIs with difference adherence level in the urban population of Ethiopia. Improving adherence to different NPIs greatly reduces *R*_*o*_ in both urban and rural population.

**Table 7:** Summary of *R*_*o*_ under the implementations of different NPIs with difference adherence level in the urban population of Ethiopia.

| Face mask | Percentage of Population who Practice Physical-distancing |  |  |  |  |  |  |  |  |  |
| --- | --- | --- | --- | --- | --- | --- | --- | --- | --- | --- |
|  | Hygiene=30% |  |  |  |  | Hygiene=40% |  |  |  |  |
|  | 5 | 10 | 20 | 30 | 40 | 5 | 10 | 20 | 30 | 40 |
| 5 | 0.7057 | 0.6686 | 0.5943 | 0.5200 | 0.4457 | 0.6049 | 0.5731 | 0.5094 | 0.4457 | 0.3820 |
| 15 | 0.6554 | 0.6209 | 0.5520 | 0.4830 | 0.4140 | 0.5618 | 0.5322 | 0.4731 | 0.4140 | 0.3548 |
| 25 | 0.6806 | 0.6448 | 0.5731 | 0.5015 | 0.4298 | 0.5834 | 0.5527 | 0.4912 | 0.4298 | 0.3684 |

### 3.6 Sensitivity Analysis

Figure 10 shows that the model is particularly sensitive to physical distancing (SD), hand wash (HW) and direct human to human disease transmission rate (*β*_1_). Hand wash and physical distancing were inversely proportional to *R*_*o*_, while *β*_1_ has directly proportional to *R*_*o*_. Parameters with relatively large PRCC values (> 0.5 or *<* −0.5) as well as corresponding small p values (*<* 0.05) are considered as the most important parameters. Parameters whose PRCC value are close to +1 or −1 most strongly influence the model. A negative sign for PRCC indicates inverse relationship of the parameter with *R*_*o*_.

**Figure 10:**
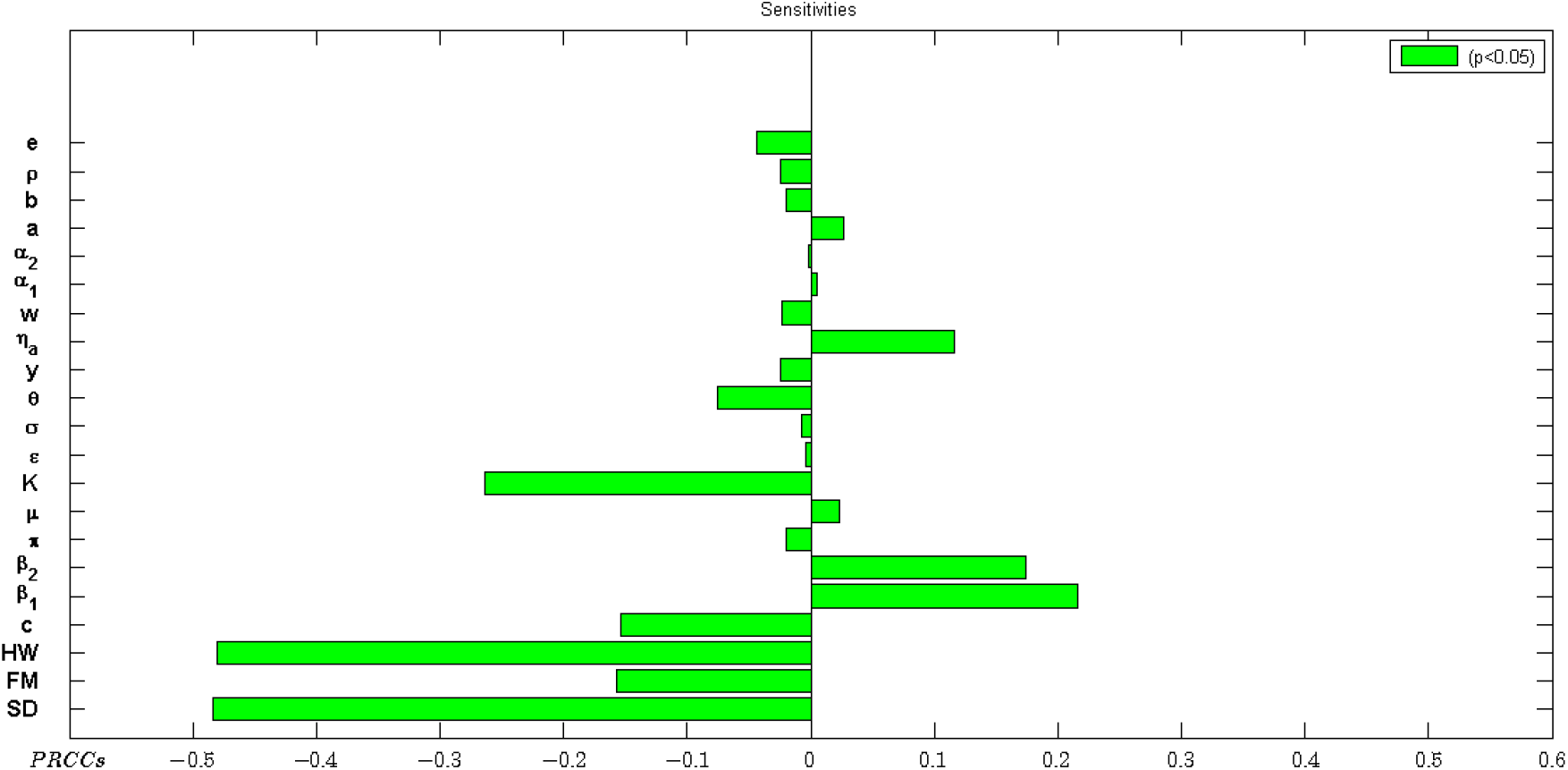
Sensitivity of parameters with respect to *R*_*o*_

To see whether one parameter depends on another, pairwise comparisons were carried out. The processes underlying the parameters physical distancing, hand washing and the proportion of the population who become symptomatic infectious, *θ*, have the greatest potential of containing the epidemic if increased, whereas processes described by *β*_1_ and *E* and *η*_*a*_ have the greatest potential of making the epidemic worse when increased (Figure 10).

In this respect, increasing physical distancing directly reduces *β*_1_ as this lowers the likelihood of a susceptible individual getting in contact with a potentially infected individual. In addition, practicing good hygiene (such as regularly washing hands, using sanitizers to disinfect the infected environment, ventilation of rooms and avoiding touching the T-zones of the face) is associated with lowering the chance of contracting the virus from infected surfaces. Anything contrary to the above increases the likelihood of getting the infection through the two aforementioned routes. Moreover, improving hand wash practices will reduce viral contamination of the environment by infected individuals.

On the other hand, the processes underlying the parameters with negative PRCC have a potential to contain the number of cases when enhanced.

To ascertain whether the process described by those parameters are different or not, a pairwise comparison to significant parameters was undertaken. Table 8 presents the computed p-values for different pairs of significant parameters by accounting for the false discovery rate (FDR). The major question posed at this point is: Are the different pairs of significant parameters different after FDR adjustment? Based on the FDR adjusted p-values in Table 8, the compared pairs of parameters are rendered to be different if their p-value is less than 0.05 and not different otherwise.

**Table 8:** Summary of different pairs of significant parameters by accounting for the false discovery rate

| | SD | FM | HW | c | $\beta_1$ | $\varepsilon$ | $\theta$ | y | $\eta_a$ | w | e |
| --- | --- | --- | --- | --- | --- | --- | --- | --- | --- | --- | --- |
| SD |  | TRUE | FALSE | TRUE | TRUE | TRUE | TRUE | TRUE | TRUE | TRUE | TRUE |
| FM |  |  | TRUE | FALSE | TRUE | TRUE | TRUE | FALSE | TRUE | TRUE | TRUE |
| HW |  |  |  | TRUE | TRUE | TRUE | TRUE | TRUE | TRUE | TRUE | TRUE |
| c |  |  |  |  | TRUE | TRUE | TRUE | FALSE | TRUE | TRUE | TRUE |
| $\beta_1$ | | | | | | TRUE | TRUE | TRUE | TRUE | TRUE | TRUE |
| $\varepsilon$ | | | | | | | TRUE | TRUE | TRUE | TRUE | FALSE |
| $\theta$ | | | | | | | | TRUE | TRUE | TRUE | TRUE |
| y |  |  |  |  |  |  |  |  | TRUE | TRUE | TRUE |
| $\eta_a$ | | | | | | | | | | TRUE | TRUE |
| w |  |  |  |  |  |  |  |  |  |  | TRUE |
| e |  |  |  |  |  |  |  |  |  |  |  |

Table 8 presents a summary of compared parameters, where “TRUE” indicates compared parameters are significantly different, and “FALSE” otherwise. The results show that more sensitive parameters are also significantly different (see Table 8) except for the pair rate of recovery of asymptomatic infectious individuals-appropriate use of face masking *y* − *FM*, recovery rate of symptomatic infectious individual-rate of going to isolation *w* − *e*, social distancing and hand washing *SD* − *HW*, respectively which may not necessarily be correlated.

## 4 Discussion and Conclusion

In this study, a modified SEIR model was developed to project the number of COVID-19 cases with different stage of the disease under the implementation of wearing face masks, hand washing and physical distancing with varying adherence levels in both urban and rural populations of Ethiopia. Unlike other mathematical modeling studies done in the context of Ethiopia Siraj et al. (2020); Alemneh and Telahun (2020); Takele (2020), this study provides projected number of all active cases, symptomatic and asymptomatic cases and ICU cases at the peak time of the pandemic. The projected number of people in each stage of the disease at the peak period and the time of the peak in urban and rural areas of Ethiopia helps the government to choose and enforce better intervention mechanisms.

Our projection shows that, in line with Howarda et al. (2020); Worby and H-H. (2020) improving the percentage of the population who wear face masks by 10% will protect around 100,000 more people from being infected by COVID-19 in the urban population. Regardless of face mask coverage and hygiene, the peak time of COVID-19 could be shifted to the future by two months if 40% of the population implements social distancing properly. Similar to our finding, the study by Siraj et al. (2020) demonstrated practicing physical distancing and wearing face masks delays the peak time of the pandemic.

In the rural population setting, implementing hygiene measures, physical distancing, and wearing face masks at 20%, 40%, and 5%, respectively, could shift the peak time into to 2021. The projected number of active cases during the peak time could reach around 360,000 if 5%, 40%, and 20% of the population wear masks, keep physical distancing and hand wash with soap, respectively. As social distancing is already a custom due to the life style in the rural population of Ethiopia, improving hygiene by 20% could help to decrease the number of cases by 2-3 fold.

In the urban population setting, if 20% of the population implements physical distancing and 30% adopt hygiene measures, and 25% wear face masks the peak time of the pandemic will happen on December, 2020 with 119,000 estimated cases (Table 3). During the peak time of the pandemic, except for some scenarios (30% physical distancing and above ≥ 15% face mask coverage), the projected number of ICU case are above the capacity of the Ethiopian health system, which needs the government attention (Table 3).

COVID-19 is a deadly virus for which vaccination or effective medical treatment is not yet available. Hence the government should focus on prevention of infection using NPI mechanisms. Low socioeconomic status and behavioral attitudes lead to closer interactions in congested households, hampering the successful implementation of social distancing measures. Lack or improper use of masks and use of low quality masks may exacerbate the spread of infection. It is therefore imperative that the prudent use of NPIs is implemented in Ethiopia to contain the pandemic. NPIs used in combination are able to decrease cases and fatalities due to Covid-19. Separate implementation of each of the NPIs shortens the peak time of the pandemic and the number of new cases will increase by two-fold. We considered the place of residence being urban and rural as a factor since adherence to the recommended NPIs and the life style and living conditions, varies greatly in urban and rural society of Ethiopia.

Similar to the urban population projection, projection of cases in the rural setting was done by modifying the model parameters in the context of rural population practices. Table 4 presented summary of projected active COVID-19 cases and ICU cases in rural population of Ethiopia under 30 scenarios. The results shows that, at the peak of the pandemic, the number of projected cases in rural population will be higher than the urban population.

## Conclusion

The first COVID-19 case was detected on March 13, 2020 and since then the government of Ethiopia has applied different mitigation strategies. But the virus continues to spread. Since community transmission of the virus is established, temporally relaxing any of the three non-pharmaceutical interventions could cause a rebound of the number of new cases, potentially leading to a collapse of the health care system. Our findings confirm, in the context of Ethiopia, that suppression of COVID-19 could be achieved by the combined implementation of three public health measures: wearing of face masks, social distancing and hand hygiene.The most effective public health measures are found to be face mask wearing in urban population and social distancing in rural populations. To mitigate COVID-19, in the urban population of Ethiopia, strict implementation of all three NPIs are required. In addition to announcing public health measures, the government should work on proper enforcement of NPIs, and the public should properly practice public health measures for their own safety and well-being. Based on the projected results under different conditions, concerned stakeholders could recommend achievable NPIs for their implementation to policy makers.

## Limitation

Studies on COVID-19 showed the distribution of cases strongly depend on age. The resources available could not get data on the number of cases by age group from concerned organizations in order to get age-structured predictions and to assess the impact of school closure on the pandemic. Further, the projection to the rural population setting is less reliable as the analysis has been constrained by different factors (i.e. exact number of incidences were not known, adherence to the implemented NPIs greatly vary across different regions of the country, estimated number of protected individuals assumed 20%,and others). Further, the study did not account for under reporting of COVID-19 cases.

## Data Availability

In this study, data on daily number of COVID-19 cases, cumulative number of deaths, and number of critical patients was extracted from Ethiopian Public Health Institute website (\url{www.ephi.gov.et/}) and Ministry of Health official Twitter page(\url{https://twitter.com/FMoHealth/}) on daily basis.

https://www.ephi.gov.et

https://twitter.com/FMoHealth/

**Appendix**

### Theorem 2.1

*Proof*. **Positivity of** Ω

To show Ω positive, first we show *S* of the model is positive for all *t* ≥ 0. To prove by contradiction: suppose, *S*(*t*) = 0, *S* (*t*) *<* 0, *E* ≥ 0, *I*_*s*_ ≥ 0, *I*_*a*_ ≥ 0, *H*_*c*_ ≥ 0, *IH*_*m*_ ≥ 0, *R* ≥ 0, *D* ≥ 0, *E*_*nv*_ ≥ 0*t* > 0. Then, using the first equation of system (1) we have,

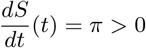

which contradicts *S′* (*t*) *<* 0. Thus, S(t) remains positive for *t* ≥ 0. For the rest of population variables we can show positivity using Gronwall’s inequality as follows.

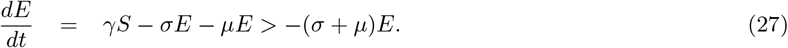

Since *S*(*t*) > 0 for *t* ≥ 0 we can use Gronwalls inequality and 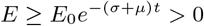.

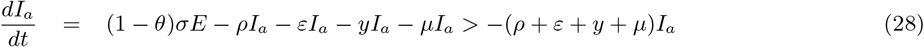

since *E*(*t*) > 0 for *t* ≥ 0 we can use Gronwalls inequality and 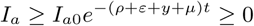.

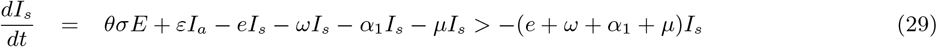

since *E*(*t*) > 0, *I*_*a*_(*t*) > 0 for *t* ≥ 0 we can use Gronwalls inequality and 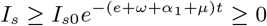.

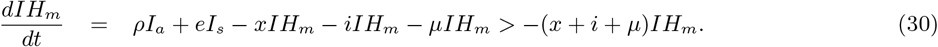

Since *I*_*a*_(*t*) > 0, *I*_*s*_(*t*) > 0 for *t* ≥ 0 we can use Gronwalls inequality and 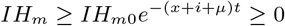.

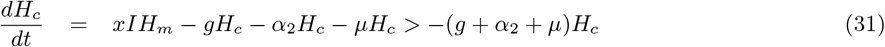

since *I*_*s*_(*t*) > 0, *IH*_*m*_(*t*) > 0 for *t* ≥ 0 we can use Gronwalls inequality and 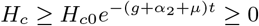.

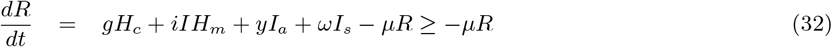

since *I*_*a*_(*t*) > 0, *I*_*s*_(*t*) > 0, *IH*_*m*_(*t*) > 0, *H*_*c*_(*t*) > 0 for *t* ≥ 0 and 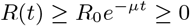.

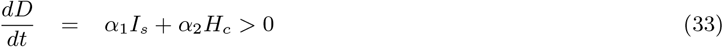

since *I*_*s*_(*t*) > 0 *H*_*c*_(*t*) > 0 for *t* ≥ 0 and 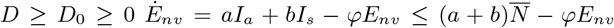. Since *I*_*a*_(*t*) and *I*_*s*_(*t*) are less than the total population *N* given as 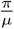 for all *t* ≥ 0. Applying again the Gronwall inequality, for 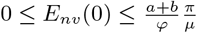 leading to

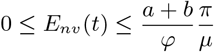

Hence all are non-negative for *t* ≥ 0. Finally, the total number of population

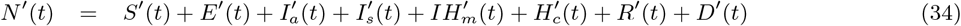

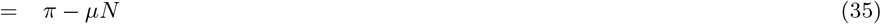

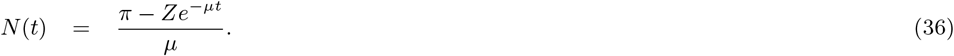

Thus for initial data 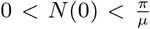 we have 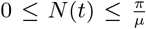. Moreover, for the environment *E*_*nv*_, we have 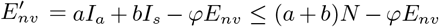 Since *I*_*a*_(*t*) and *I*_*s*_(*t*) are less than *N* for all *t* ≥ 0 applying Gronwalls inequality for 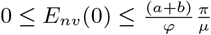 gives 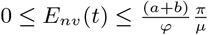.

## Footnotes

2 COVID-19 ICU:32.9% died, 44.1% recovered https://www.bbc.com/news/uk-scotland-52653192

